# No magic bullet: limiting in-school transmission in the face of variable SARS-CoV-2 viral loads

**DOI:** 10.1101/2022.03.25.22272956

**Authors:** Debra Van Egeren, Madison Stoddard, Abir Malakar, Debayan Ghosh, Antu Acharya, Sk Mainuddin, Biswajit Majumdar, Deborah Luo, Ryan Nolan, Diane Joseph-McCarthy, Laura F. White, Natasha S. Hochberg, Saikat Basu, Arijit Chakravarty

## Abstract

In the face of a long-running pandemic, understanding the drivers of ongoing SARS-CoV-2 transmission is crucial for the rational management of COVID-19 disease burden. Keeping schools open has emerged as a vital societal imperative during the pandemic, but in-school transmission of SARS-CoV-2 can contribute to further prolonging the pandemic. In this context, the role of schools in driving SARS-CoV-2 transmission acquires critical importance. Here we model in-school transmission from first principles to investigate the effectiveness of layered mitigation strategies on limiting in-school spread. We examine the effect of masks and air quality (ventilation, filtration and ionizers) on steady-state viral load in classrooms, as well as on the number of particles inhaled by an uninfected person. The effectiveness of these measures in limiting viral transmission is assessed for variants with different levels of mean viral load (Wuhan, Delta, Omicron). Our results suggest that a layered mitigation strategy can be used effectively to limit in-school transmission, with certain limitations. First, poorly designed strategies (insufficient ventilation, no masks, staying open under high levels of community transmission) will permit in-school spread even if some level of mitigation is ostensibly present. Second, for viral variants that are sufficiently contagious, it may be difficult to construct any set of interventions capable of blocking transmission once an infected individual is present, underscoring the importance of other measures. Our findings provide several practical recommendations: the use of a layered mitigation strategy that is designed to limit transmission, with other measures such as frequent surveillance testing and smaller class sizes (such as by offering remote schooling options to those who prefer it) as needed.

## Introduction

Continued high levels of SARS-CoV-2 transmission strain healthcare systems and accelerate viral evolution, which undermines vaccinal efficacy^1,2^, and generates new variants with unpredictable epidemiological characteristics. For example, the transmissibility of SARS-CoV-2 has increased over time, with the Delta variant being around twice as transmissible^3^ as the original Wuhan strain, which had an *R*_0_ (reproduction number) of between 3.3^4^ and 5.7^5^. The incubation time of the disease has also demonstrated evolutionary change, going from 5-21 days for the original Wuhan strain^6,7^ to 2-4 days for the Omicron variant^8^.

In order to slow viral evolution by limiting transmission, it is thus important to understand the role of schools in facilitating SARS-CoV-2 spread. In most countries, a significant fraction of the population consists of K-12 students, staff and first-degree household contacts of students and staff. In the US, 40% of all households have a child at home under 18 years of age^9^, and 23% of the US population is enrolled in school^10^. Thus, in-school transmission of SARS-CoV-2 will substantially impact transmission dynamics in the whole population.

A number of studies during the early part of the pandemic led to the perception that SARS-CoV-2 did not spread in schools, based on the similarity in case counts between schools and their surrounding communities and a lack of observed transmission chains among children in schools. However, the methodological validity of these conclusions is debatable, as the metrics being used to infer a lack of spread are themselves vulnerable to a “false negative” problem (absence of evidence is not evidence of absence) (see Supplementary Text S2, and ^11^ for a detailed critique on the limitations of current research arguing that SARS-CoV-2 does not spread in schools). In fact, there is now a robust body of evidence supporting the contention that SARS-CoV-2 spreads efficiently in schools that lack adequate infection control measures. Empirical analyses using county-level panel data in the United States have demonstrated that counties with fully open K– 12 schools with in-person learning had a 5% increase in the growth rate of COVID-19 cases during April-December 2020 (a period of time when US schools were largely closed for the first five months)^12^. Consistent with this finding, COVID-19 symptom reporting was more common in areas where schools were open compared to areas with remote learning, an effect that was attenuated in communities using multiple mitigation measures^13^. In-school transmission is apparent when systematic surveillance testing methods are used^14–17^, and dramatic increases in case detection rates have also been observed in studies that relied on surveillance as opposed to symptomatic testing^18^.

A layered mitigation strategy is one way to limit transmission in the school setting under conditions of widespread community transmission. There are multiple potential interventions available at this point: vaccines, masks, air quality improvements, surveillance testing, contact tracing/ isolation and podding. For reasons of cost and practicality, it is rational to seek a minimal set of infection-control measures. The challenge in this regard is that ongoing viral evolution can yield further changes to the characteristics of the virus, and a set of infection control measures that works well for one viral variant may readily be defeated by the next.

An important open question then is: “What is the design of a minimal set of infection-control measures in schools that is robust to variant-to-variant differences in viral load?” In this work, we use mathematical modeling of the steps involved in viral transmission to understand the impact of infection-control measures in a range of different scenarios corresponding to variants with differing viral loads. Our intent was to address both the feasibility and robustness of strategies for limiting SARS-CoV-2 transmission in schools.

## Results

To study SARS-CoV-2 transmission and the impact of control measures in schools, we created a multistep mathematical model of viral transmission in indoor settings (Methods) occurring under the assumption of indoor aerosol spread of infectious virus (see Supplementary Text S3 for justification of this assumption). First, we estimated the concentration of virions in the air over time in a room with an infected individual present using a differential equations model. This model assumes that infected individuals emit virions into the air at a constant rate into a room that is modeled as a well-mixed container (see Supplementary Text S4 for details about this assumption). Emitted virions can be inactivated over time or filtered out via air exchanges. These viral concentration estimates were then used as an input to calculate the probability that uninfected individuals in the room will become infected with SARS-CoV-2.

### Viral concentrations in a room with an infected individual present reach steady state quickly

Under this model of SARS-CoV-2 emission, the viral concentration in a room reaches a steady-state concentration after the infected individual has been present for a certain time. For a closed room without air exchange or filtration, this concentration *C*_SS_ depends on the emission rate *α*, viral inactivation rate *δ*, and the volume of the room *V* and is estimated as *C*_*SS*_ = *α*/(*δV*) (Methods). To understand the impact of evolutionary changes in viral load, we picked three variants of the virus whose viral loads have been reported in the literature to be widely different from one another (see Supplementary Text S5 for details about viral loads for the Wuhan strain and the Delta and Omicron variants). For a typical classroom size (20 feet x 20 feet x 10 feet), the steady-state concentration with an individual infected with the original Wuhan virus is approximately 4 virions per liter, which scales linearly for higher viral load SARS-CoV-2 strains (Fig. 1). We found that this steady-state concentration is reached after an infected individual is present in a room for 1-2 hours, regardless of the rate at which the individual emits viral particles. Masking of the emitter decreases the rate at which infected individuals release virions, lowering the final steady-state concentration, but does not affect the time at which the steady state is reached (see Supplementary Text S6 for details about parameter estimates for various interventions in this and subsequent sections). We further explore the effects of different types of masks on the steady-state viral concentration later in this section (Fig. 6). After the infected individual leaves the room, the viral concentration returns to approximately zero over a 5-hour period in a closed room (Fig. S1).

**Figure 1:**
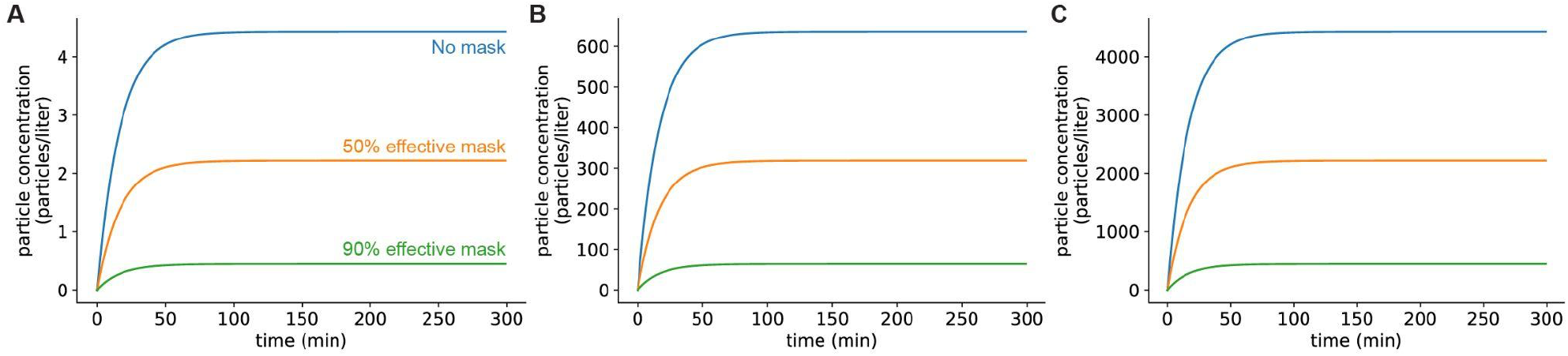
Viral concentrations in a room with an infected individual present reach steady state quickly. **A:** Infected individual with a low viral load strain of virus (similar to the initial Wuhan strain). **B:** Individual infected with an intermediate viral load variant (similar to Omicron). **C:** Individual infected with a high viral load variant (similar to Delta). Room dimensions are 10’ x 20’ x 20’. No air filtration or other mitigation methods were used. In all panels, the blue curve shows the concentration when the infected individual has no mask on, the orange curve shows the concentration when the infected individual has on a mask which filters out 50% of exhaled particles (typical cloth mask), and the green curve shows the concentration when the infected individual has on a mask which filters out 90% of exhaled particles (typical surgical mask).

### Steady-state concentrations of virions are reduced by high-volume air filtration

Filtering the air in the room decreases the steady-state concentration of virions by increasing the rate at which infectious virions are eliminated from the room (Methods). For infected individuals with a low viral emission rate (e.g., infected with the original Wuhan SARS-CoV-2 virus), air filtration and masking of the infectious individual can reduce the steady-state concentration of virus in the room to less than one virion per liter of air (Fig. 2). However, for SARS-CoV-2 variants and individuals with higher viral loads and therefore higher viral emission rates (e.g., with Omicron-like viral loads, represented here as the ‘medium’ rate, or with Delta-like viral loads, represented here as the ‘high’ emission rate), the steady-state viral concentration in the room might remain high even with high filtration efficiency and filtration rate (Fig. 2). Notably, for schools using high efficiency filtration systems (e.g., HEPA filters), increasing the rate of air exchange across the filter is important for minimizing transmission.

**Figure 2:**
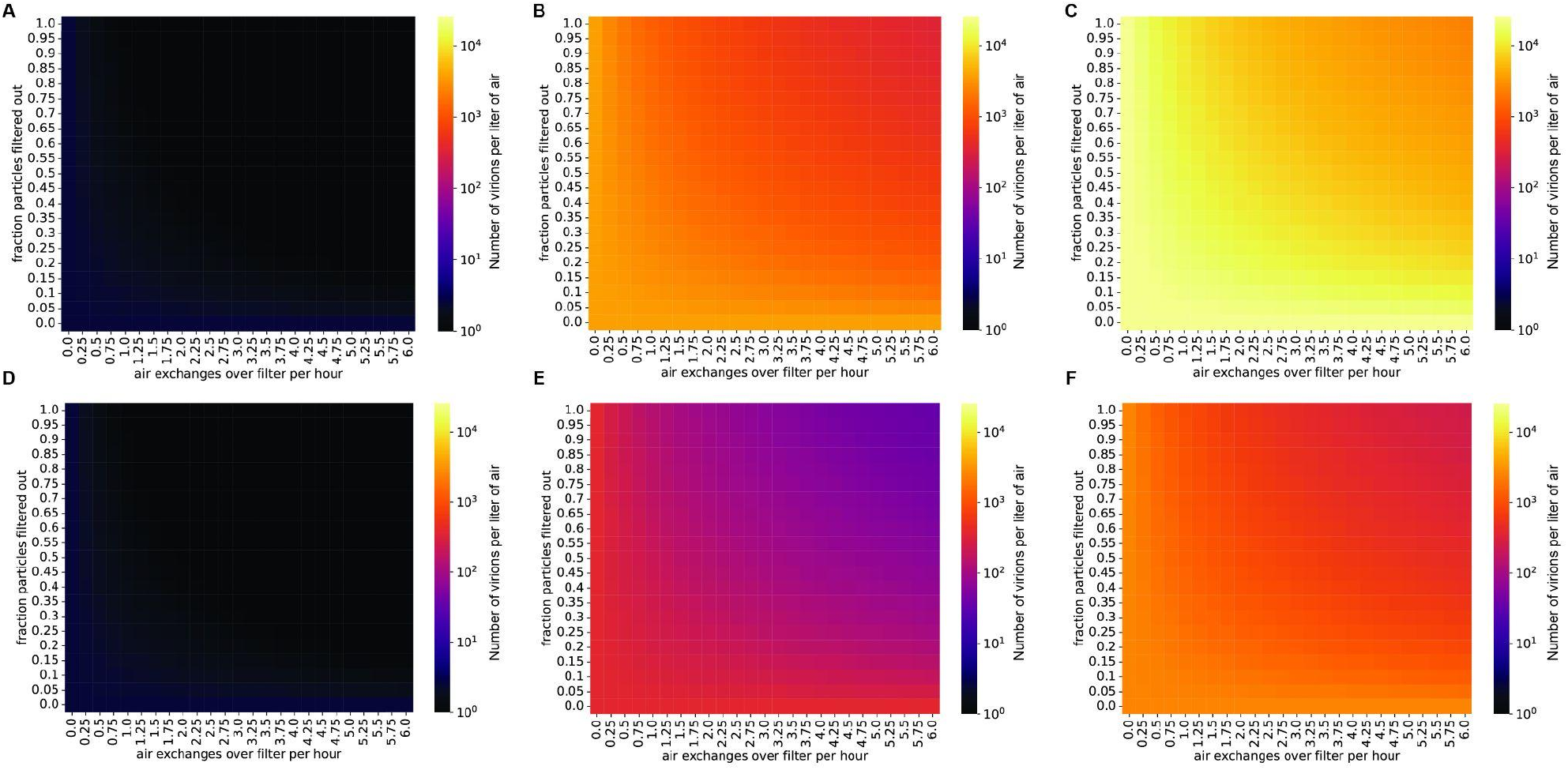
Steady-state concentrations of virions are strongly impacted by air exchange rate across the filter and by the fraction of particles removed when air is passed over the filter. Virion concentrations at steady state, in a room 10’ x 20’ x 20’. **A-C:** Individual infected with a low viral load (A), intermediate viral load (B), or high viral load (C) strain, no mask. **D-F:** Individual infected with a low viral load (D), intermediate viral load (E), or high viral load (F) strain, wearing a 90% effective mask. Typical air purification systems can achieve a filtration rate of approximately 5-6 exchanges/hr (Supplementary Text S6).

### Steady-state concentrations of virions in the air are decreased by ionizers

Another strategy to lower the viral concentration is to use ionizers, which inactivate viral particles and remove them from the air^19^. These devices produce small ions by the corona discharge principle, according to which negatively charged ions transfer their charge to suspended particles upon collision. These charged particles then agglutinate, becoming larger until they fall out of the air under the effect of gravity^20^. Ionizers generating negatively charged ions have been shown to be efficient at removing bacteria, molds, and viruses from indoor air^21–23^. The efficiency of particle removal is dependent on the emission rate of ions within an enclosed space, as well as room volume^24^. Studies conducted with smoke particles in an enclosed room suggest a high efficiency of ionizers in removing particles from the air, that varies between 80 and 100%^20,24,25^. Although older ionizer technologies generate ozone, which is an undesirable byproduct, newer ionizers do not have this potential liability associated with them^26^.

As with the other control measures we simulated, we found that ionizers can lower viral concentrations in a typical classroom to below one virion per liter in situations where the infected individual is emitting viral particles at a relatively low rate (masked, infected with a low viral load strain) (Fig. 3). However, if the individual is infected with a high viral load SARS-CoV-2 variant (e.g., Delta), the viral concentration in the room will be very high even when ionizers are being used (Fig. 3).

**Figure 3:**
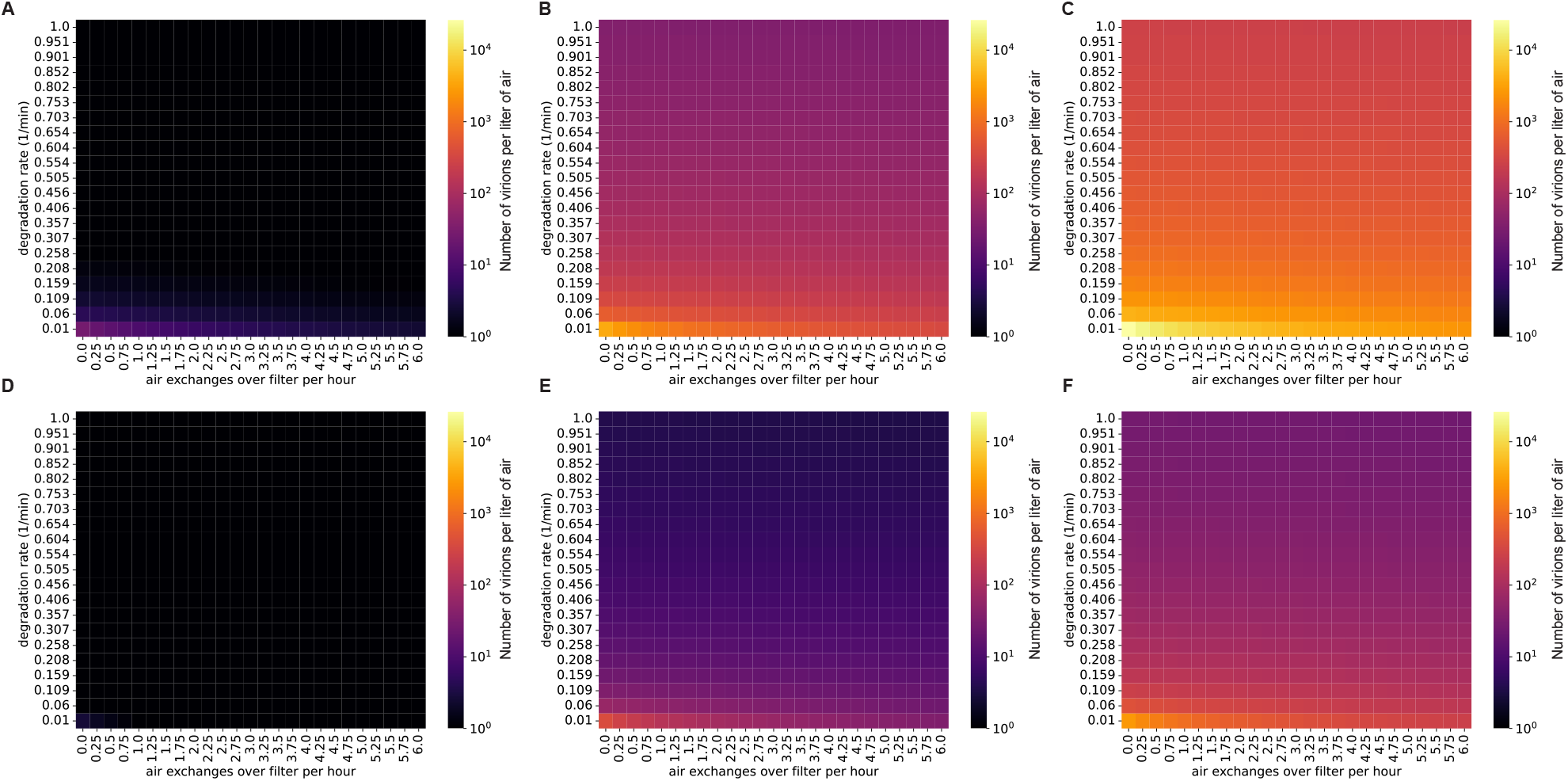
Steady-state concentrations are decreased by ionizers (which increase the particle degradation rate). Virion concentrations at steady state, in a room with the same dimensions as Figure 1. **A-C:** Individual infected with a low viral load (A), intermediate viral load (B), or high viral load (C) strain, no mask. **D-F:** Individual infected with a low viral load (D), intermediate viral load (E), or high viral load (F) strain, wearing a 90% effective mask. The degradation rate of airborne SARS-CoV-2 without the use of an ionizer is approximately 0.01/min.

### Only a fraction of inhaled viruses is deposited in the nasopharynx

We used the viral concentration estimates calculated in the previous section, estimates of aerosolized particle deposition derived from computational fluid dynamics (CFD) modeling, and the minimum infectious dose of SARS-CoV-2 to estimate the probability that uninfected individuals in the room will become infected with SARS-CoV-2.

Infection of a new host requires the virus to be inhaled and deposited on the airway mucosa, where it can replicate. To study transmission dynamics in indoor settings with airborne SARS-CoV-2, we used a published computational fluid dynamics model of airflow in the nasopharynx to estimate the number of inhaled viral particles that are deposited in the airway mucosa^27^. This computational model of airflow in the human nasopharynx estimates the probability that an inhaled virion will reach the airway mucosa, given the size of the liquid droplet in which it is suspended. We assumed individuals breathe in virions suspended in liquid droplets with the measured steady-state size distribution of expelled respiratory droplets^28^. We used the computational fluid dynamics modeling results to compute the overall probability that an inhaled virion will hit the nasopharynx, marginalized over the empirical droplet size distribution (Methods, Table S1), and found that approximately 0.6% of virions that are inhaled during each breath are deposited in the mucosa.

### Masks and air filtration reduce the expected number of virions transmitted to uninfected individuals

We used this nasopharyngeal deposition probability to estimate the number of virions that reach the airway mucosa per hour in an uninfected student in a classroom with a given concentration of SARS-CoV-2 in the air (Methods). For a classroom with an individual emitting virions at a low rate, interventions such as masking and air filtration can lower the transmission rate to an uninfected individual to less than 1 virion per hour, preventing transmission for short periods in the classroom (Fig. 4). However, exposure over several hours or an entire school day may transmit enough virions to the nasopharynx to cause infection, so limiting contact and masking of uninfected students is still important for controlling viral spread in this situation. If the virus-emitting individual is infected with a high viral load variant (similar to Delta), the number of virions inhaled can increase to hundreds or thousands per hour, possibly causing infection even after only a short exposure period (Fig. 4). In fact, while several hours are required for transmission of a low viral load (Wuhan-like) variant in a classroom, an individual infected with a high viral load (Delta-like) variant can spread the virus within a matter of minutes (Fig. 5). Thus, in this scenario, with multiple environmental control measures (masking and air filtration) in place, transmission can still occur over an 8-hour school day. Thus, in a future scenario where we are faced with a Delta-like variant with high viral loads, schools may have to rely on additional measures (e.g., rapid and widespread surveillance testing, targeted school closures) to limit in-school spread. These measures would need to supplement and not replace in-school mitigation measures in order to be effective.

**Figure 4:**
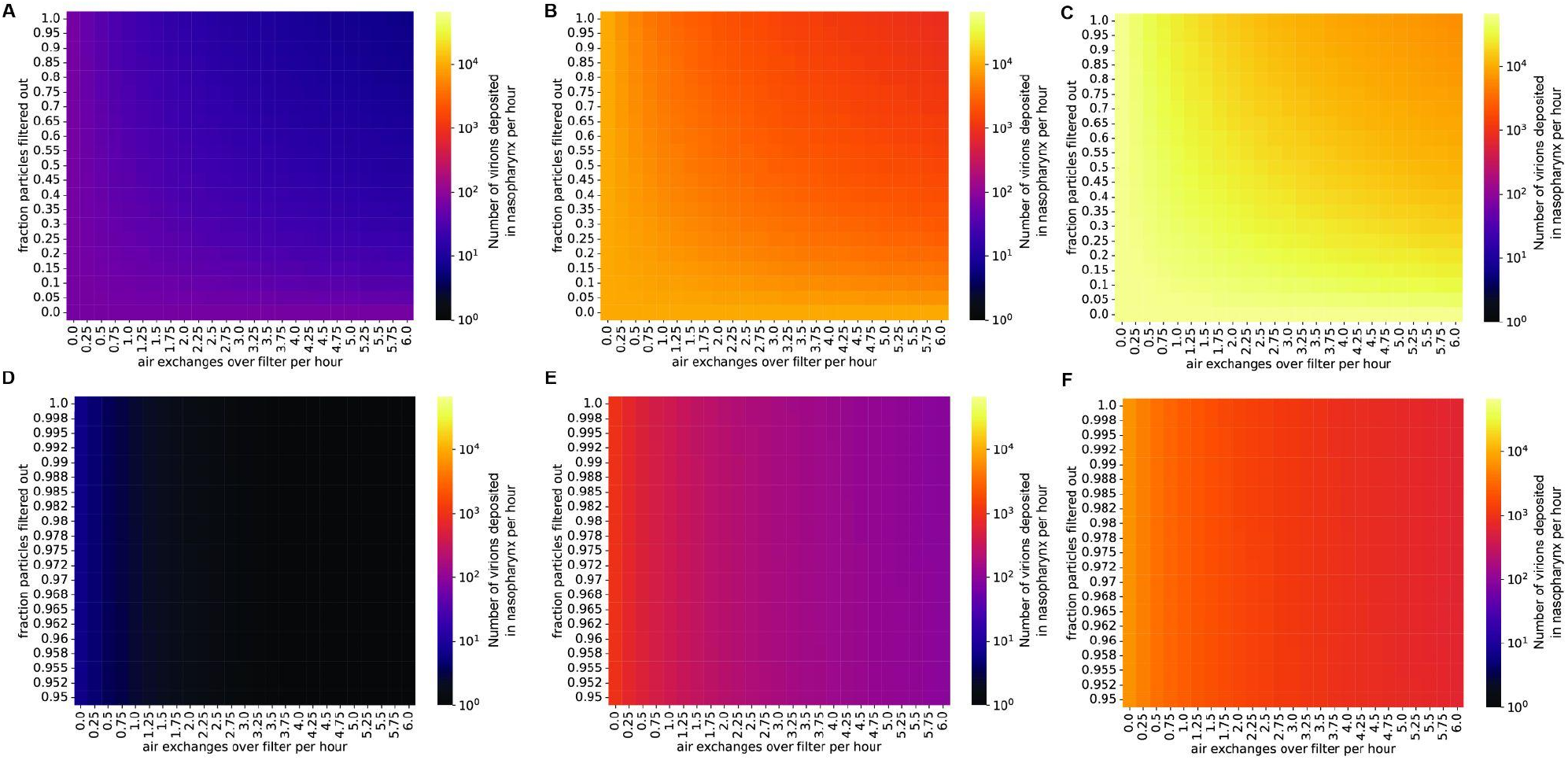
Even with masks and air filtration, a person exposed to a high viral load emitter will be exposed to thousands of virions per day. Virions inhaled by an uninfected individual per hour in the presence of an infected individual, in a room with the same dimensions as Figure 1. The probability that an inhaled virion hits nasopharynx was estimated at 0.6% (Methods). **A-C:** Individual infected with a low viral load (A), intermediate viral load (B), or high viral load (C) strain, no mask. **D-F:** Individual infected with a low viral load (D), intermediate viral load (E), or high viral load (F) strain, with the emitter wearing a 90% effective mask (one-way masking).

**Figure 5:**
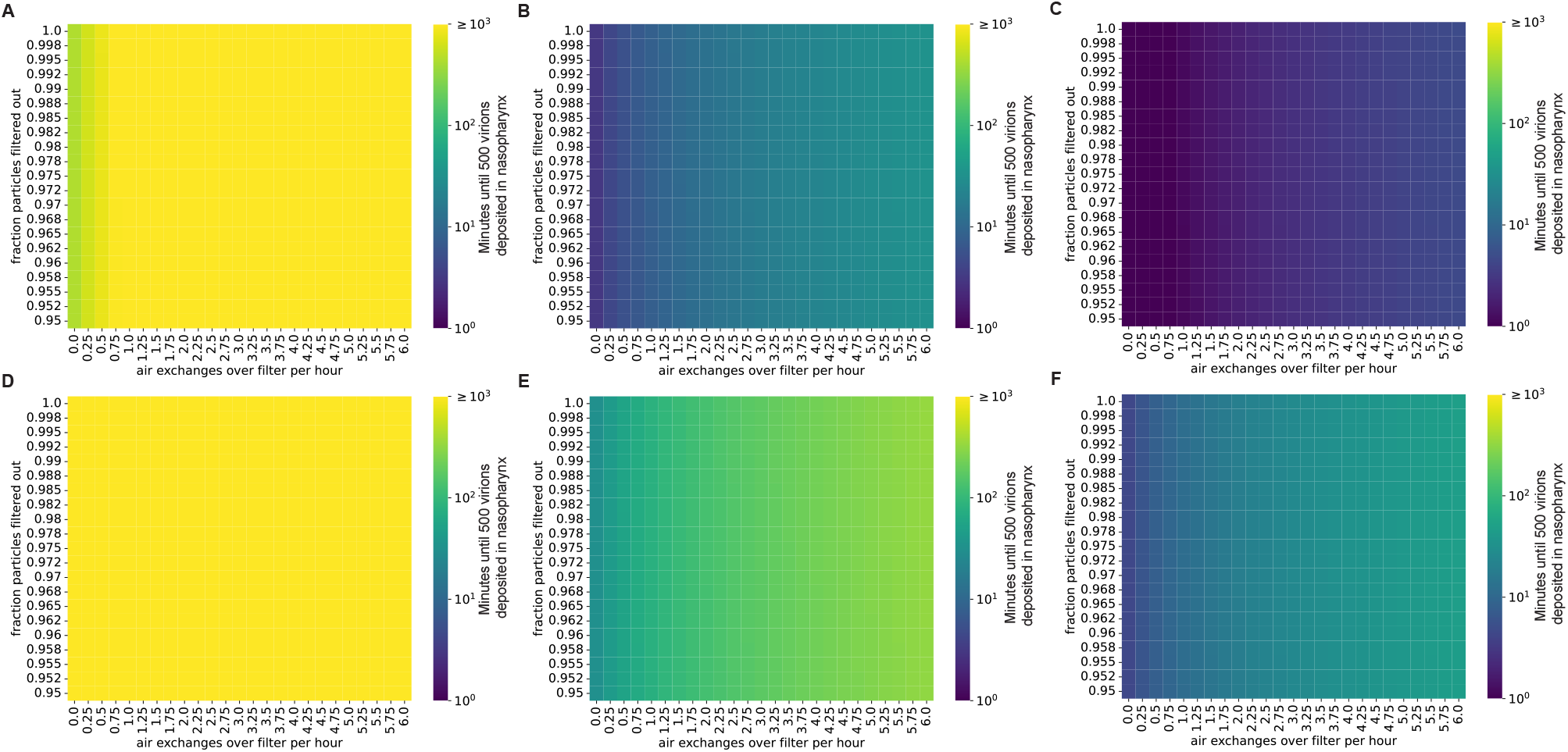
The presence of robust mitigation measures substantially increases the time to infection for low viral load strains. Plots show the time until an uninfected individual receives 500 virions in a room occupied by an infected individual. **A-C:** Individual infected with a low viral load (A), intermediate viral load (B), or high viral load (C) strain, no mask. **D-F:** Individual infected with a low viral load (D), intermediate viral load (E), or high viral load (F) strain, with the emitter wearing a 90% effective mask (one-way masking).

### Two-way masking can substantially reduce the number of virions transmitted

High-quality masks, used correctly, have been demonstrated to reduce infection risk even in high-risk settings^29–31^. Therefore, such masks can provide an additional level of mitigation in the event that schools are faced with a high viral load (Delta-like) variant. Masks provide a double benefit, as they reduce transmission by filtering out virions emitted by infected individuals and by reducing the number of ambient virions inhaled by uninfected individuals. Masks that filter out >95% of virions increase the time to transmission by approximately 10-fold when worn on either the infected or uninfected individual (one-way masking; Fig. 6). If both the infected and uninfected individuals are masked (two-way masking), less-effective masks (e.g., well-fitted surgical masks) can achieve the same level of protection against transmission. Combining universal N95 masking with excellent ventilation can increase the time to transmission of even high viral load strains to longer than a typical school day (Fig. 6D), suggesting that layered mitigation strategies featuring well-fitted and high-quality masks are critical for the control of high viral load strains in the classroom.

**Figure 6:**
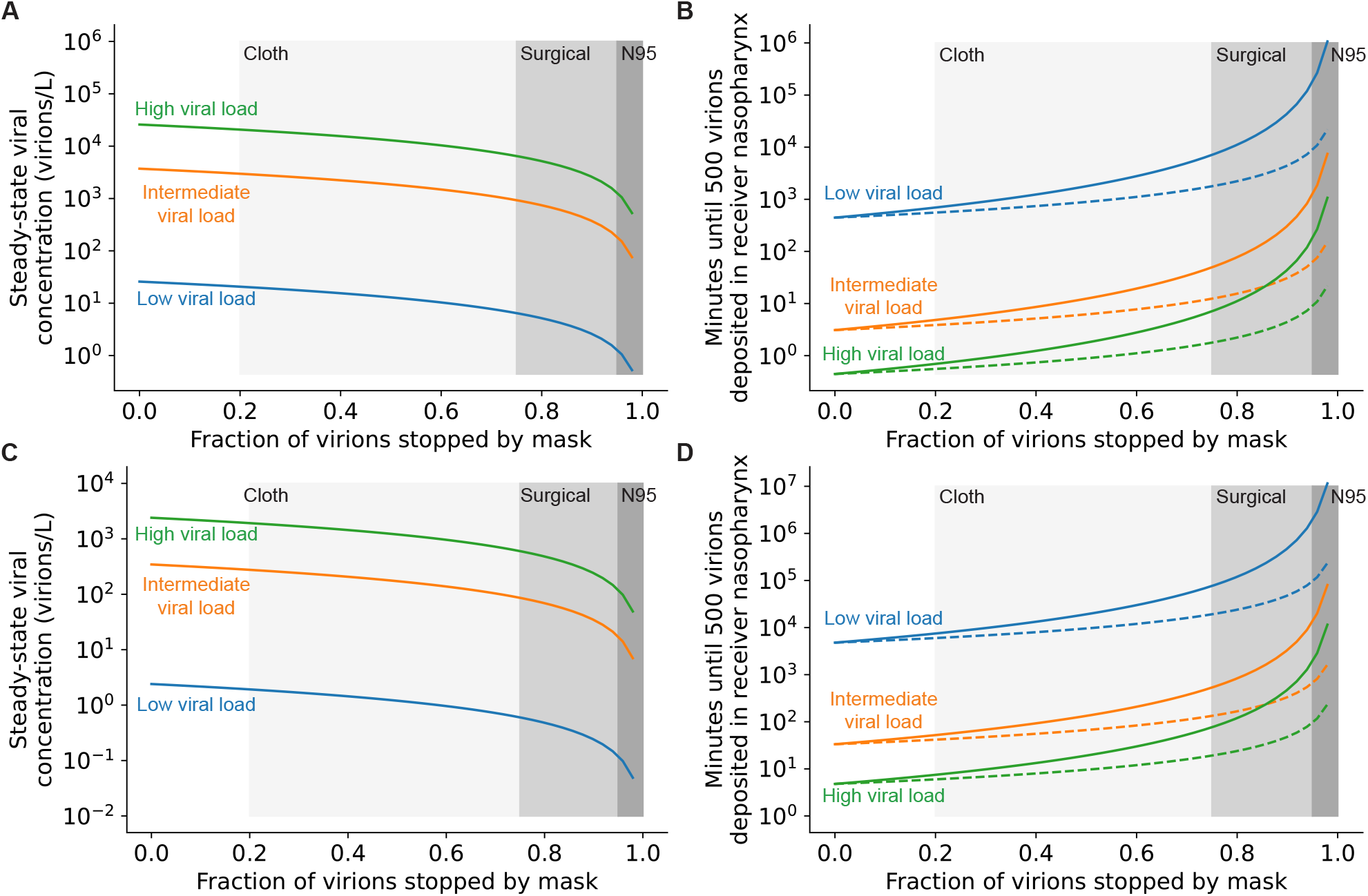
Masking both infected and uninfected individuals reduces the rate of viral transmission. **A:** Steady-state airborne viral concentration in an unventilated classroom with a single infected individual wearing a mask with the filtration efficiency given on the x-axis. **B:** Time until infection of uninfected individuals present in an unventilated classroom with a single infected individual wearing a mask with the filtration efficiency given on the x-axis. Uninfected individuals either are not wearing masks (one-way masking; dashed lines) or are wearing a mask with the same filtration efficiency as the infected individual (two-way masking; solid lines). **C:** Steady-state airborne viral concentration in a well-ventilated classroom (6 complete air exchanges per hour) with a single infected individual wearing a mask with the filtration efficiency given on the x-axis. **D:** Time until infection of uninfected individuals present in a well-ventilated classroom with a single infected individual wearing a mask with the filtration efficiency given on the x-axis. Uninfected individuals either are not wearing masks (one-way masking; dashed lines) or are wearing a mask with the same filtration efficiency as the infected individual (two-way masking; solid lines). In all panels, shaded regions denote typical filtration efficiencies for cloth, surgical, and N95 masks (Supplementary Text S6) and curve colors denote viral load of the emitter.

### In a well-ventilated room, risk is strongly dependent on seating position relative to the infected individual

Up to this point, we have considered the classroom to be a well-mixed container. This simple modeling approach allows us to identify settings where the risk of in-school transmission is high. The well-mixed container assumption is justified, particularly in settings with limited ventilation (see Supplementary Text S4 for details). However, in some settings, the assumption may not hold, particularly in well-ventilated rooms, which are thought to have a low risk of transmission overall. To better understand the risk of transmission in a well-ventilated setting, we used CFD simulations of air transport inside a classroom. To simulate a best-case scenario for ventilation, we chose to model airflow behavior with the dimensions of a large auditorium-style classroom in a tropical setting, with high ceilings and several windows that are all open (Fig. 7A-C). We placed a single infected individual (emitter) in the room and varied their position to understand the impact of the airflow in the room.

**Figure 7:**
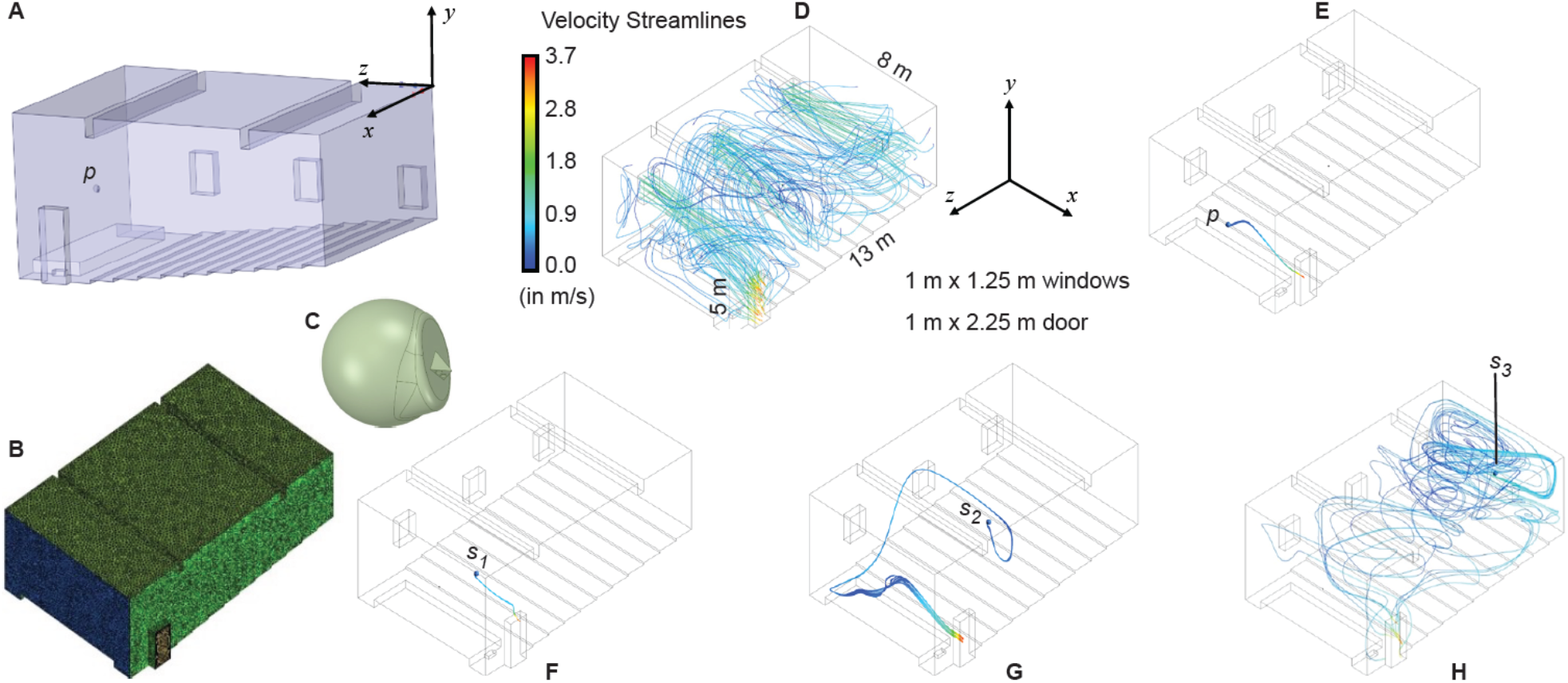
In a well-ventilated room, infection risk is strongly dependent on the location of the infected individual. **A:** Digitized geometry of a classroom (based on direct measurements of an auditorium-style classroom in a tropical setting). The position of the instructor’s head is marked by *p*. **B:** Meshed space interior to the bounds of the classroom. **C:** Typical head geometry for the emitter subtracted from the classroom mesh, with realistic nostril diameters. **D**: Mixing of velocity streamlines in the classroom with the windows as the inlet and the door as the outlet. The panel includes the layout and plan dimensions of the ventilated classroom. **E-H:** Flowlines of exhaled virions showing the impact of emitter location (blue dot: p represents the instructor’s head location, *s*_*1-3*_ represent the students’ head locations) on local concentrations of virions. Note that *s*_*1*_ is in the front seat, *s*_*2*_ is in the middle of the classroom, and *s*_*3*_ is in the rear seat. Local concentrations are dependent on fluid dynamics within the classroom, suggesting that even if the average concentration of virions in a room is below the infectious threshold, individuals may become infected over time based on their location downstream of the emitter.

Figure 7D visually depicts the airflow mixing trends inside the room and the virion-bearing streamline patterns for emitters located at different parts of the auditorium-style classroom. As the room is well-ventilated, the aerosolized virions emitted by the teacher (Fig. 7E) standing on the podium at the front of the room (with a door located proximally on the side), as well as those from the student seated in the front row (Fig. 7F), would escape through the door quite readily. The situation is, however, different if the infected individual were seated in the middle of the room (Fig. 7G), and even worse if the infected individual were seated at the rear (Fig. 7H). In these two situations, the infected individual would be efficiently spreading aerosolized pathogens, via exhaled respiratory ejecta, through the entire room. Thus, for a well-ventilated room (where the well-mixed container assumption cannot be expected to apply) total viral load in the room depends strongly on the position of the infected individual in the room. Additionally, local virion concentrations may be sufficiently high to enable efficient viral spread in the absence of other countermeasures (such as masking). The chaotic airflow patterns (invisible to the naked eye) underscore the unpredictable downside of infection risk in a closed setting (Fig. 7H).

Thus, while it is possible to use model-based approaches to identify settings with a high risk of transmission, model-based approaches that rely on the well-mixed container assumption cannot definitively identify indoor settings with a low risk of transmission. This finding further underscores the need for multiple layers of intervention, and a robust ability to detect outbreaks before they spread.

### Measures to reduce the likelihood of infected individuals being present in the classroom setting are crucial

Environmental control measures (masking, air filtration, ionizers) can all have an impact on limiting transmission in the in-school setting. However, our work suggests that these measures-both individually and in concert-are all vulnerable to defeat by a sufficiently high burden of virion emission. Thus, reducing the probability of infected individuals being present in the classroom at all is crucial to limiting SARS-CoV-2 transmission in schools. To that end, three measures are critical for reducing the expected number of infected individuals in a classroom: testing, capacity limits, and targeted school closures. Regular screening tests, with infected individuals being identified and isolated before they enter the classroom, can reduce the number of expected infected individuals arriving at school each day. Reducing class sizes by running the school day in shifts or by offering a remote school option to students that prefer it can also reduce the expected number of infected individuals in schools (Fig. 8). Podding, which limits students’ exposure to others outside of their cohort, such as in the lunchroom, can interrupt transmission chains to prevent spread throughout the school. Similarly, keeping schools closed for periods of time when local transmission is high would also have a proportional impact on reducing transmission. Notably, widespread vaccine coverage -- which is essential to limit the mortality and morbidity burden of the ongoing COVID-19 pandemic--may not play a significant role in limiting transmission at present (see Supplementary Text S8 for a summary of vaccinal efficacy against transmission).

**Figure 8:**
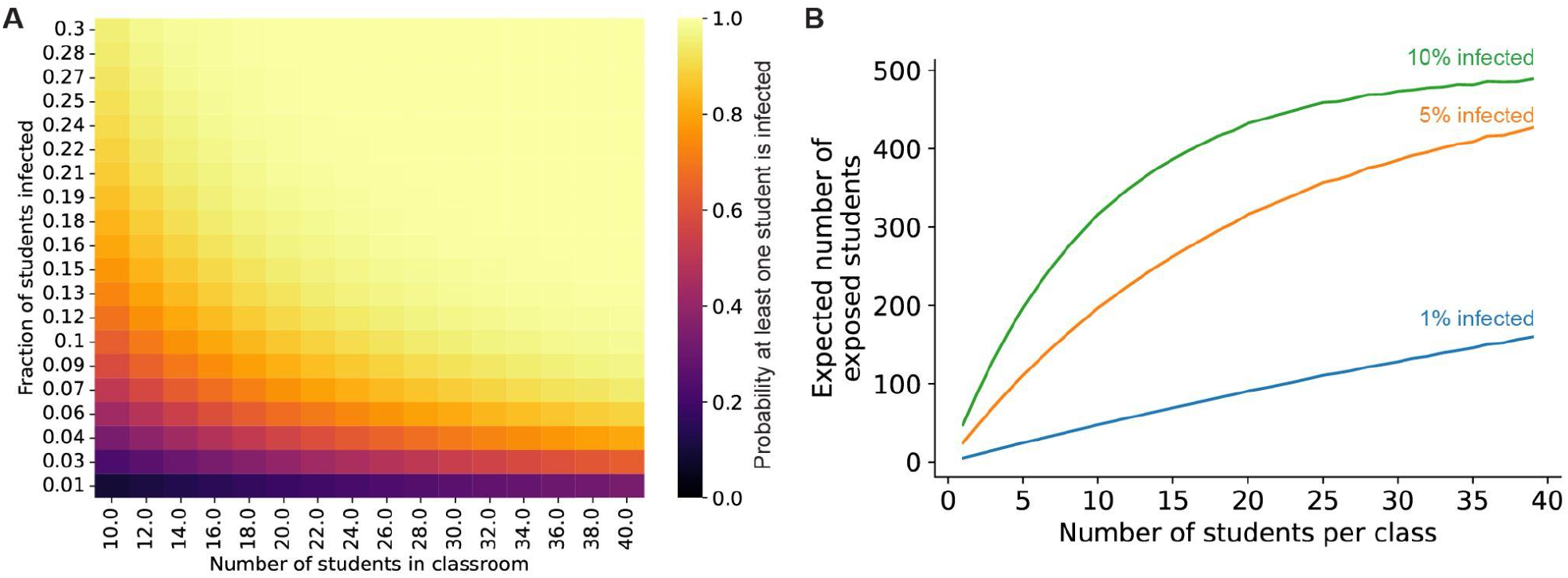
Reducing class sizes reduces the risk of infection in school settings. **A:** Probability that at least one student in a classroom is infected, given values for the population-level infection prevalence (y-axis) and the number of students in the classroom (x-axis). **B:** Expected number of students exposed to an infected individual in a 500-person school when the school is broken up into different size classes over the entire school day (see Methods for additional details), given different values for the initial overall SARS-CoV-2 infection prevalence in the student population (blue curve: 1% of students infected, orange curve: 5% infected, green curve: 10% infected).

## Discussion

In this study, we have used mathematical modeling to demonstrate the strengths and limitations of a layered mitigation strategy in limiting in-school transmission while keeping schools open. We examined the impact of risk mitigation measures (masking, ionizers, ventilation, filtration) on limiting spread within a classroom when an infected person is present. Our findings underscore the critical importance of layered mitigation strategies in limiting in-school transmission. With that said, all of the examined measures can be readily defeated by sufficiently high viral loads, a biological change that has already been observed during the pandemic (for example between the Wuhan strain and the Delta variant). This is a crucial point: minimal effective measures for the disease as it is at present may have an increased risk of failure in the face of new variants of SARS-CoV-2. Our findings also indicate that the risk of transmission in schools may be hard to predict in certain settings (such as in the turbulent airflow patterns of a well-ventilated room). As a corollary, our work points to the central importance of relying on measures to limit the likelihood of having an infected person in the classroom: testing and isolation, limiting class sizes, and targeted closures when community transmission is high.

The ongoing COVID-19 pandemic shows no signs of permitting a return to pre-pandemic life, with high rates of transmission leading to rapid viral evolution. The course of the pandemic under these conditions is likely to be unpredictable, with the potential for catastrophic levels of mortality and morbidity should new variants of the virus emerge that have greater immune evasion potential or virulence. Thus, slowing viral evolution by limiting transmission is now a vital societal imperative.

There are many examples in human society where the routine operation of vital services provided by complex systems brings some measure of risk to those involved: road transportation, aviation, medical care, law enforcement, agriculture and power generation, to name a few. In each of these cases, risks are managed by using a systems approach^32^. The premise is that accidents in complex systems occur through the accumulation of failures. This “Swiss Cheese model”^33^ of risk mitigation is well known in the epidemiological community, and a number of epidemiologists have advocated for its use from the beginning of this pandemic^34,35^. The strength of the Swiss Cheese model in risk management lies in building a system that is robust to human error. In the specific case of an evolving virus, the Swiss Cheese model also provides multiple orthogonal selection pressures, making escape more difficult for the virus. To keep schools open, designing guidelines to limit transmission based on the Swiss Cheese model, and then validating those guidelines to ensure robustness to epidemiological changes driven by viral evolution, will be critical.

In the school setting, the application of the Swiss Cheese model has been slow for a number of reasons: the risk of in-school transmission has been under-estimated or compared to the wrong outcome (health effects on children), and false dichotomies in strategic thinking have led to an inappropriate focus on some layers of the “Swiss Cheese” in preference to others (for example, arguing for unmasking because children are vaccinated, or arguing for eliminating testing and contact tracing because children are masked). At this point, there is clear evidence supporting the contention that SARS-CoV-2 transmission can occur in schools^11,36^. The public-health consequences of that transmission are not borne by children alone. In addition to the first-order effect of household transmission of school-acquired COVID-19, transmission in the school setting facilitates viral evolution. An evolving virus benefits from a narrow focus on individual measures-the more focused the selection pressure, the easier it is for the virus to escape it. The mistake of over-reliance on one layer of the Swiss Cheese Strategy (made with the vaccines) can easily be repeated with other measures (such as testing and contact tracing).

This work has a number of assumptions and key limitations. We have assumed for most of the work that the classroom is a well-mixed container, and then demonstrated (Figure 7) that the failure of this assumption leads to higher risk than could be estimated from a well-mixed container assumption. We also assume that children are equally susceptible and infectious as adults (see Supplementary Text S1 for an in-depth discussion of this assumption). It assumes perfect compliance with mask-wearing, which is not likely to be true in practice^37,38^. We also have not considered the effect of vaccination on transmission or risk of infection-primarily because the role of vaccines in limiting SARS-CoV-2 infection (and transmission) has now been demonstrated to be highly time-sensitive and vulnerable to immune evasion (See Supplemental Materials S8 for an in-depth justification of this assumption).

There are a number of very thoughtful modeling analyses on this topic that have been published throughout the course of this pandemic. Several other groups have used model-based analyses to demonstrate that in-school transmission of SARS-CoV-2 is likely to be significant^12,13,39^. The point that reopening schools without robust COVID-19 mitigation could lead to an acceleration of the pandemic has been made in several modeling studies^40–42^. At the same time, a number of groups have published models focused on a limited set of infection-control measures, to show how schools can be reopened without risking in-school transmission (for example by using portable air purifiers^43^, limiting class sizes^44^, or mandating vaccination^45,46^). Our work adds to the discussion by pointing out the need to take evolutionary-driven epidemiological changes (for example due to viral load from one variant to the next) into account. In his book *The Black Swan* (now a classic in the risk-management community), author Nasim Nicholas Taleb argues that the key to risk management lies not so much in predicting the worst thing that could happen, but in making plans that are robust to that outcome.

Our modeling demonstrates that a layered mitigation strategy, implemented properly, can curtail viral transmission under many circumstances. With that said, there are ways to implement infection-control measures that are ineffective, and measures that are effective in the presence of one viral variant can be readily rendered ineffective in the presence of another. Because many of the interventions have a nonlinear effect on risk mitigation, cutting corners on risk mitigation steps can degrade their utility very quickly, turning them into “hygiene theater”. For example, we found that two-way masking with N95 masks increases the time until SARS-CoV-2 transmission by multiple orders of magnitude, as compared to one-way masking with cloth or surgical masks (Fig. 6). As a corollary, it is crucial for schools to have controls in place to ensure that measures taken for mitigation are working as intended. For example, air quality can be monitored using carbon dioxide monitors, and mandatory (as opposed to opt-in) testing can be used to monitor the functional outcomes of in-school mitigation. Mitigation strategies should be pressure-tested using simple mathematical modeling approaches such as the one described in this paper. Thresholds for the acceptable performance of mitigation measures (for example, air filters or ionizers) should be updated periodically to reflect changes in viral epidemiology. Thus, only by iterative optimization of control measures can we expect to have effective suppression of in-school transmission of SARS-CoV-2. Taking a risk-management mindset to the problem of developing layered measures for infection control in the school setting is crucial at this stage. Optimizing such approaches to ensure robustness in the face of viral evolution may allow us to escape the false dichotomy of keeping schools open versus bringing the pandemic to an end.

## Methods

To estimate the risk of viral transmission we built a multistep model comprising emission of viral particles into the air and subsequent inhalation and deposition into the nasopharynx of uninfected individuals (Fig. S2).

### Estimating viral steady-state concentrations in a room with an infected individual

We designed a mathematical model of viral concentrations in a room in which an infected individual emits airborne SARS-CoV-2 particles at a constant rate. We assumed that the airborne virions are distributed uniformly throughout the room (i.e., the air in the room is well-mixed). Virions are inactivated at a constant rate and have an average lifetime of approximately 1.6 hours indoors at 73°F, 55% humidity^47^. For a closed room with no air exchange or filtration, the change in virion concentration over time is given by the differential equation

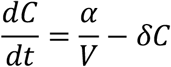

where *α* is the viral emission rate, *δ* is the viral inactivation rate, *V* is the volume of the room, and *t* is the amount of time elapsed since the infected individual entered the room.

Therefore, the viral concentration in the air over time *C*(*t*) is

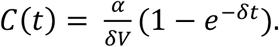

Air filtration removes virions in the room by filtering out a certain fraction of viruses that pass through the filter. Assuming the room is well-mixed, this adds an additional first-order virus removal process with rate *εβ*/*V* where *ε* is the fraction of virions that are eliminated while passing through the filter (between 0 and 1), *β* is the rate at which room air is passed through the filter (in units of air exchanges per hour), and *V* is the room volume. With filtration, the concentration in the air is

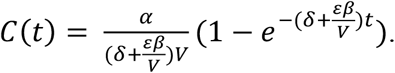

Ionizers simply increase the virus inactivation rate *δ*.

For the original Wuhan virus strain, we estimated the rate at which infected individuals emit viral particles into the air using published data which show that rooms with infected individuals have approximately 2-6 viral copies per L of air^48^. Assuming these measurements were taken when the virus was at a steady-state concentration in the room and that these individuals were in a reasonably sized room (4000 cubic feet of air), we estimated that the viral emission rate would be approximately 2319-6937 virions/h using our equation for the viral steady-state contribution shown in the Results. Therefore, we used a value within this range (5000 virions/h) for the emission rate for the low viral load simulations in this study. See Supplemental Materials for details about parameter estimates for various interventions.

### Estimating the risk of infection based on the steady-state viral concentration in a room

To cause a new infection, virions must be inhaled by an uninfected host and be deposited in the airway mucosa, where they can replicate and cause disease. The rate at which virions are inhaled is the product of the respiratory tidal volume (volume of air inhaled during each breath) and the respiratory rate. However, computational fluid dynamics suggests that a minority of inhaled virions hit the nasopharyngeal mucosa, and this probability depends on the size of the liquid droplets containing the virions^27^. We used published estimates of the respiratory droplet size distribution^28^ and the probability of hitting the nasopharynx for different inhaled droplet sizes^27^ to estimate the overall fraction of inhaled virions that are deposited in the airway mucosa, marginalized over the empirical respiratory droplet size distribution, as

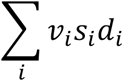

where *s*_*i*_ is the fraction of exhaled droplets of size *i, d*_*i*_ is the probability of mucosal deposition for droplets of size *i*, and *v*_*i*_ is the fraction of aerosolized virions that are contained in droplets of size *i*. This fraction *v*_*i*_ is calculated as

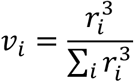

where *r*_*i*_ is the radius of the droplets of size *i*. To calculate the time until infection of an uninfected host, we conservatively assumed that 500 virions were required for infection (see Supplementary Text S7 for details about the minimum infectious dose).

### Numerical simulations of air mixing and exhalation patterns inside a realistic ventilated classroom

Flow physics play a vital role in the distribution of airborne pathogens inside a confined space, e.g., a classroom. To model that, we have implemented state-of-the-art Computational Fluid Dynamics (CFD) simulations of: (a) air transport inside the room owing to incoming flux through the open windows; and (b) exhaled streamline patterns from human emitters situated at different points of the room. The digital reconstruction of the room is based on the measurements from an auditorium-style classroom. The room is sized at 13 m × 8 m × 5 m, with three windows (each 1 m × 1.25 m) and a door (1 m × 2.25 m). A head-sized sphere is used to stand in for the emitter’s head in each simulated flow model and is subtracted from the corresponding interior mesh. The sphere is provided with nasal protuberances, bearing nostrils realistically sized at 107.65 mm^2^ and 125.33 mm^2^, based on computed tomography reconstructions of human subjects^27^. For the emitter’s locations, we tested four different scenarios: (i) streamlines emitted by the teacher standing on a podium at the front of the classroom, (ii) a student seated in the front row, (iii) a student seated in the middle of the auditorium, and (iv) a student situated at the rear row. The classroom interior is meshed on ICEM CFD 2019 R3 (ANSYS Inc., Canonsburg, Pennsylvania) with tetrahedral elements bearing a maximum size of 0.15 m. The resolution was based on an earlier study^49^ on meshing requirements (maximum size < 0.2 m) to numerically model natural ventilation. Additionally, five layers of prism elements^49^ with a height ratio of 1.2 are extruded on the boundary walls. The meshed geometries are then imported to ANSYS Fluent 2019 R3 to track the air transport using the SST k-*ω*model^50^ to account for the turbulent flow scales.

It is noted that when wind encounters a blocking effect on its path owing to a building, the velocity pressure is converted into static pressure. Consequently, on the windward side, the pressure would increase, with consequent pressure reduction on the leeward side. The static pressure gradient generated by wind on the building surface can be estimated by:

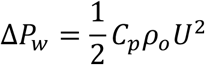

where Δ*P*_*w*_ is the static pressure difference, *C*_*p*_ is the wind pressure coefficient, *ρ*_*o*_ is the density of outside air, and *U* is the mean wind velocity. The *C*_*p*_ value, which is independent of wind speed but depends upon wind direction and incident angle, can be calculated for low storied buildings (for up to 3 stories)^51^. The other physical parameters, namely *ρ*_*o*_ and *U* are taken as 1.139 Kg/*m*^2^ and 2.7 m/s, from earlier studies^52^ and meteorological data^53^, respectively. Air dynamic viscosity was assumed to be 1.9065 × 10^−5^ kg/m.

However, there will be another pressure difference – one between the classroom and the outside corridor (next to the door). From established building code standards^54^, we can conclude that pressure differential Δ*P*_*indoor*_ between the classroom and corridor will be 5 – 20 Pa, resulting in the net pressure gradient from window-to-door (i.e., main inlet to main outlet) to be

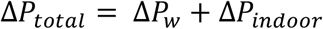

From the above assessments, the individual pressure gradients approximate to Δ*P*_*w*_ = 3.5 Pa and Δ*P*_*indoor*_ = 5 Pa. In the simulations: windows were taken as pressure inlets with 0-gauge pressure, nostrils were taken as velocity inlets with volumetric flux at 15 L/min^27,55–58^ to replicate gentle steady breathing, and at the door, we imposed a pressure outlet condition with negative gauge pressure of Δ*P*_*total*_, i.e., -8.5 Pa. Wall boundary condition was mimicked as a stationary wall with no slip condition.

### Modeling impact of class size on SARS-CoV-2 exposure

The probability that at least one student in a classroom arrives infected with SARS-CoV-2 was estimated from the overall prevalence of the infection using Poisson statistics. This probability *P* was estimated as

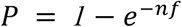

where *n* is the number of students in the classroom and *f* is the fraction of infected individuals in the overall population. To estimate the expected number of exposed individuals in a school with *N* total students, the entire student population was broken into classes with *i* students each. The probability that at least one student in each class *P*_*i*_ was estimated using the above equation, and the expected number of students exposed to an infected individual was then calculated as *NP*_*i*_.

### Data and materials availability

Scripts implementing the viral concentration and transmission models are available on GitHub at https://github.com/dvanegeren/covid-indoor-transmission. Input data used to estimate the fraction of inhaled droplets that hit the nasopharynx are also available in the same GitHub repository.

## Data Availability

Airborne viral concentration simulation data are reproducible using the code available at https://github.com/dvanegeren/covid-indoor-transmission. All additional data are available from the authors upon reasonable request.

