## Supplementary Text S1-S8, Figs. S1 and S2, Table S1 for "No magic bullet: limiting in-school transmission in the face of variable SARS-CoV-2 viral loads"

#### S1: Children's susceptibility and infectiousness

Early in the pandemic, there was a perception that children had reduced susceptibility to infection by SARS-CoV-2, based on studies conducted while children had lower contact rates than the general population[1,2]. Rigorous meta-analyses do not support children's reduced susceptibility to infection at this point[1]. Consistent with this, studies with surveillance testing strategies report similar rates of infection between school-aged children and adults. For instance, in the UK (October 2020), the highest prevalence of SARS-CoV-2 infections in areas with open schools was in 18-25 year-olds, followed by 11-18 year-olds, with 5-11 year-olds having comparable prevalence to working-age adults[3]. At present, within the United States, the American Academy of Pediatrics[4] estimates that greater than 1 in 10 children in the country has tested positive, and children comprised 19.0% of the cumulative reported COVID-19 cases (22.2% of the US population is under the age of 18).

The efficiency with which children transmit SARS-CoV-2 infections has also been debated- a key finding reported and cited often in the early debate about school reopening was that children were not usually the index case (first infection) within in a family[5], suggesting that children may not be responsible for disease spread. However, this finding is confounded with the lower likelihood of detection of asymptomatic index cases, and a number of studies (many of which rely on surveillance testing) point to children's infectivity being similar[6–10] or even higher[11,12] than that of adults. Notably, similar infectivity between children and adults has been reported for variants of concern such as B.1.1.7[13].

#### S2: Methodological issues with inferences regarding lack of transmission in school settings

The CDC's science brief on the topic of in-school SARS-CoV-2 spread states that "the majority of cases that are acquired in the community and are brought into a school setting result in limited spread inside schools when multiple layered prevention strategies are in place”[14]. This inference is based on two metrics.

The first metric is that rates of infection in schools and communities usually track each other closely, as has been reported in numerous studies worldwide. This correlation has been used inappropriately by the CDC and others[15,16] to infer a lack of a causal relationship. In prior work[17], we examined the validity of this inference by simulating a scenario where schools have a higher rate of SARS-CoV-2 transmission than the surrounding community and comparing it to a scenario where the rate of transmission was similar between schools and the community. We found that the ratio of cases between schools and community tracked closely even when schools were driving spread. This finding suggests that the correlation between caseloads in schools and communities is just as likely to be a result of in-school spread driving transmission within communities (cannot use correlation in case counts to infer a lack of causality). In prior work, we have demonstrated that transmission chains originating within schools are capable of generating large chains of spread within the community that can remain undetected in the absence of widespread surveillance testing[18].

The second metric supporting the idea of limited spread in a school setting is that chains of transmission that can be clearly linked to in-school disease spread are rare. However, absence of evidence is not evidence of absence. Symptom-gated forward contact tracing (the method commonly used by schools in the US) to detect child-to-child transmission relies on the appearance and reporting of two consecutive symptomatic cases, connected by a transmission event (for example, see [16]). Because the majority of transmission comes from a minority of cases (overdispersion)[19] and children are more likely to experience asymptomatic infections than adults[20], symptom-gated forward contact tracing is expected to detect only 4.4% of all child-to-child transmission events in schools[17].

In fact, this may point to a problem with symptom-gated forward contact tracing in general- recent contact-tracing studies in the United States suggest that many named contacts are not successfully traced[21,22] and not all symptomatic contacts are willing to undergo testing[23]. Consistent with this, other events which may plausibly have led to rampant disease spread were not shown to have done so by symptom-gated forward contact tracing. As an example, consider the case of the Sturgis motorcycle rally in August 2020, a 10-day event in Meade County, South Dakota attended by approximately 460,000 persons without any mask-wearing requirements or other mitigating policies. The event was followed by a wave of COVID-19 cases in Meade County and South Dakota in the month following the rally, and counties outside of South Dakota that contributed the highest inflows of rally attendees experienced a 6.4-12.5% increase in COVID-19 cases relative to counties without inflows[24]. Despite clear evidence of population-level changes in COVID-19 case counts in the weeks following the rally, the CDC and Minnesota Department of Health were able to identify only 21 person-to-person transmission events[25]. The methodology used for contact tracing was again, voluntary symptom-gated contact tracing. Out of the 86 positive cases, only 41 reported being in close contact (defined as being within 6 feet of another person for ≥15 minutes) with other people, and they reported an average of 2.5 close contacts. Both statistics are implausible for a 10-day motorcycle rally featuring indoor dining and concerts[26–28]. The CDC's report does not specify how many of the 102 secondary contacts were tested- this is also typical for contact-tracing studies in the United States[21,22].

Taken together, this suggests that the rarity of transmission chains in a given setting should be interpreted with caution if the methodology of contact tracing is not transparent, and if voluntary symptom-gated contact tracing methods are used.

**S3: Evidence supporting the modeling assumption of aerosol spread of SARS-CoV-2**

At this point, a robust body of evidence supports the assumption that SARS-CoV-2 spread occurs primarily through aerosol transmission.

First, efficient indoor transmission is more consistent with aerosol spread than it is with other modes of spread (such as ballistic droplets or surface transmission). In this context, the transmission rate for SARS-CoV-2 has been reported to be many (~19-40) times higher indoors than it is outdoors[29,30]. There is also direct evidence for long-range transmission indoors[31,32], even in cases where people were in adjacent rooms[33] or in rooms separated by a corridor[34] despite never have been in each other's presence. Modeling suggests that long-range transmission may also occur outdoors[35].

Second, infectious virus has been isolated from a number of locations that are consistent with aerosol spread. A number of groups have reported direct isolation of infectious virus from the air[36–38], in air filters and building ducts[39], as well as in exhaled aerosols[40].

Third, epidemiological characteristics of SARS-CoV-2 are consistent with aerosol transmission. For example, aerosol spread provides a direct mechanistic basis for superspreader transmission[41], which has been well documented for SARS-CoV-2[19]. Asymptomatic transmission from people who are not coughing or sneezing – another extensively documented feature of SARS-CoV-2 - is also consistent with aerosol transmission[42,43].

There are a number of excellent overviews on the topic of aerosol spread of SARS-CoV-2[44,45]. The data supporting aerosol transmission of SARS-CoV-2 was also put forth in an open letter to the WHO[46], which the WHO initially contested in a scientific brief[47] before grudgingly accepting[48]. It is worth noting that the propensity for aerosol transmission seems to be impacted by evolution as well- for example, the Alpha variant of SARS-CoV-2 has been experimentally shown to be better able to spread via aerosol transmission than the ancestral strain[49].

**S4: Evidence supporting the modeling assumption of the room as a well-mixed container**

In this work we started with the well-mixed assumption for air flow. This initial assumption is justified from two different lines of evidence.

*Physics-based considerations*: The current state of evidence suggests a very strong contribution of aerosol particles to the spread of SARS-CoV-2 (summarized in the preceding section). In the context of this work, we have considered particles to be aerosolized if their diameters are in the range of 0.1-30 microns in diameter. For particles in this size range, the rate of spread due to diffusion or Brownian motion is negligible relative to the rate of spread due to air flow. The spread of such particles emitted by an unmasked individual due to speaking[50], coughing or sneezing[51] would be primarily expected to occur due to the horizontal momentum of exhaled air particles, with a minimal contribution from gravity-induced drift[52]. However, in the presence of masks, the horizontal movement of exhaled air is greatly suppressed, and instead the particles rise due to turbulent buoyant airflow[53]. The Reynolds number (*Re*) determines the behavior of air flow with a given speed and length scale, with moderately high *Re*s (>100) associated with vortex shedding, and still higher *Re*s (>2000) associated with turbulent flow. In the presence of forced convection within a room (such as can be expected from air circulation or ventilation), *Re* can be expected to be fairly high (≈2000), corresponding to a mix of vortex shedding and turbulent flows[54]. Additional factors can also contribute to turbulence, such as human movement.

*Epidemiological considerations:* A number of epidemiological studies also support the well-mixed-container assumption. Indoor transmission has been demonstrated to occur in a wide variety of settings at ranges that were likely greater than 6 feet apart[31,32,55]. A notable example of this was the Skagit Valley Chorale superspreader event, where a weekly rehearsal with 61 masked attendees led to 53 infections– a model-based analysis of this event has argued that the outcome supports the well-mixed assumption empirically[42]. Also consistent with the well-mixed assumption is the finding that transmission occurs 19 times more efficiently indoors[29].

The well-mixed assumption is implemented in our modeling using the Wells-Riley approach, first proposed by Wells[56] in 1955 and extended by Riley[57] in 1978. This approach has been found to be broadly applicable for indoor air transmission for other infectious respiratory diseases[58], and has been used to model SARS-CoV-2 transmission by a number of other groups[59].

### S5: Estimates for variation in viral load between viral variants and individuals infected with SARS-CoV-2

In this study, we have used the Delta variant as an example of a SARS-CoV-2 viral strain with increased viral load, to exemplify the impact of viral load on guidelines for preventing transmission. The increased transmission rate of the Delta strain relative to prior SARS-CoV-2 variants has been at least partly attributed to a higher viral load in the nasopharynx. Studies have found RT-qPCR cycle threshold values for Delta that are approximately 6-1000 times higher than prior variants. We note that there is a range of estimates for relative viral load for the Delta variant compared to other variants (see table below).

Wide person-to-person variation in exhaled viral load have also been reported for SARS-CoV-2 infections based on intrinsic factors as well as on the specific activity being undertaken[60].

| **Relative viral load of Delta** | **Variant compared** | **Reference** |
|---|---|---|
| 6.06 times greater | Alpha | [61] |
| 11.48 times greater | Pre-Alpha | [62] |
| 1260 times greater | Pre-Alpha | [63] |

Incubation times also vary between different SARS-CoV-2 variants. The incubation period for the original Wuhan strain is around 5 days[64,65] while the Delta incubation is shorter (4.3 days[64]). The Omicron variant has an even shorter incubation period, estimated at approximately 3 days[66].

**S6: Parameters governing the efficacy of individual control measures**

Air filtration:

High-efficiency particulate air (HEPA) filters are defined by the US Environmental Protection Agency to remove at least 99.97% of airborne particles for all particle sizes[67]. These filters can be installed in small mobile or large floor-standing air purifiers, which can ideally exchange 5 to 6 times the air volume in the room per hour[68,69]. A standard mobile air purification setup in a classroom has been shown experimentally to reduce airborne viral concentrations by 90% within approximately 30 minutes[69].

Ionizers:

Bipolar air ionizers create an electrostatic charge on airborne particles, causing them to be removed from the air by increasing their aggregation rate and deposition rate on surfaces[70]. Smoke particle studies suggest that ionizers can remove between 80-100% of particles from room air[71–73].

Masks:

Face masks have been shown to lower the risk of SARS-CoV-2 transmission[74] by both reducing the number of virions emitted by infected individuals and reducing the amount of virus inhaled by uninfected individuals who are masked. Cloth and surgical masks have typical filtration efficiencies of around 20-95% for droplets or aerosols, with cloth masks having lower efficiencies (20-75% particles filtered out) for smaller particles sizes (0.3-0.5um)[75]. N95 and KN95 respirators are rated to remove 95% or more of 0.3um particles[76].

**S7: Estimates for minimum infectious dose for SARS-CoV-2**

For the ancestral Wuhan strain of SARS-CoV-2, estimates of infectious dose have been made by diverse methods including CFD modeling[77] and phylogenetic analysis[78]. These estimates point to a small number of infectious particles - 6[78] to 300[77] - being sufficient to start an infection. The upper end of this range is similar to the infectious dose for SARS-CoV[79], and an order of magnitude lower than that of influenza[80]. Here, we used 500 virions as the minimum infectious dose, which is on the upper end of the published range. Estimates of infectious dose for novel variants such as Delta have been a further order of magnitude lower, with some reports suggesting that fewer than ten viral particles may be sufficient to start an infection[63,81,82].

**S8: Performance of vaccines in limiting transmission of SARS-CoV-2**

Vaccines against SARS-CoV-2 currently possess high levels of efficacy in preventing severe disease and death from COVID-19[83] and form a critical last line of defense in the public-health strategy. Vaccinal efficacy against severe disease appears somewhat stable over time, which some have suggested is linked to T-cell activity[84].

The impact of vaccines in limiting infection and transmission appears to be dependent on humoral immunity[85], and this impact appears more limited and time-dependent. Estimating the impact of SARS-CoV-2 vaccines on transmission ($VE_t$) is challenging, because many vaccine trials did not directly assess vaccine efficacy against transmission, focusing instead on symptomatic infections. As a substantial portion of SARS-CoV-2 infected patients are infectious while asymptomatic[86] or presymptomatic[87] and this proportion rises with breakthrough infections[88–90], estimates for vaccinal efficacy against transmission are biased when considering only symptomatic infections in the denominator[91,92]. Despite this overestimation bias for $VE_t$, recent reports point to a very low degree of vaccinal efficacy against symptomatic disease in some circumstances[93]. This suggests that $VE_t$ is even lower and may in some settings be negligible.

Humoral immunity wanes shortly after vaccination, as neutralizing antibodies (nAbs) decline rapidly after the second dose[94,95], with a mean half-life of about four months[96]. Consistent with this decline in nAb levels, waning vaccinal immunity against infection has been documented extensively, with substantial loss of protection against infection occurring within the first six months[97,98]. While this efficacy against symptomatic infection is restored by booster doses, booster efficacy also declines rapidly[93].

Viral evolution is a second contributing factor to the loss of vaccinal immunity. Viral immune evasion has also been demonstrated to potently reduce the ability of neutralizing antibodies (nAbs) to bind SARS-CoV-2 spike protein - for example, the Omicron variant shows a profound (20-40 fold) reduction in the binding potency of nAbs against the viral spike protein[99–102]. Consistent with this, vaccinal efficacy against symptomatic infection with Omicron is severely compromised- for example, for individuals vaccinated with two doses of the Pfizer vaccine, VE against infection is 8.8% (95% CI, 7.0 to 10.5) at 25 or more weeks[93]. Other viral variants have also demonstrated substantial reductions in nAb binding potency[103–105] and vaccinal immunity against infection[106,107]. A number of studies have pointed to a predictive relationship between nAb binding potency and vaccinal protection against infection[85,96].

In addition, the impact of vaccination on onward transmission by infected individuals has also been found to be modest. Some reports from earlier in the pandemic indicated a 50% reduction in infectiousness associated with vaccine breakthrough cases with the Wuhan strain[108]. However, the reduction in viral load for breakthrough cases is minimal for recent variants such as Delta[85,109–111] and Omicron[112]. For these variants, epidemiological data is also consistent with a picture of efficient transmission by breakthrough cases- the secondary attack rate for infection resulting from a vaccinal breakthrough case is only marginally lower than that of unvaccinated individuals for Delta[109,110] and Omicron[113]. Multiple real-world examples of superspreader events[114] and ongoing spread[115] among highly vaccinated populations add further weight to the inference of limited impact of vaccination on curbing SARS-CoV-2 spread. Going forward, continued viral evolution and waning vaccinal effectiveness[116] in reducing viral load (which has now been noted for the booster dose as well[117,118] can be further expected to impact the vaccinal reduction of transmission.

In summary, these findings support the assumption that vaccination status does not contribute significantly to the ability of an individual to contract or transmit SARS-CoV-2 in a congregate setting.

### Supplementary Figures

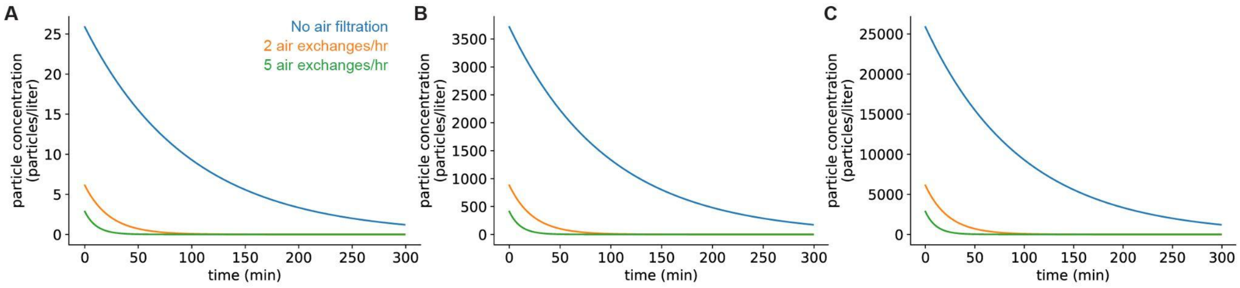


**Figure S1:** Viral concentrations in a room after the infected individual leaves. The concentration of SARS-CoV-2 in room air is initially at steady state. **A:** Infected individual with Wuhan strain of virus, **B:** Individual infected with Omicron variant. **C:** Individual infected with Delta variant. In all panels, the blue curve shows the concentration when there is no ventilation in the room, the orange curve shows the concentration when there is 2 air exchanges/hr, and the green curve shows the concentration when there is 6 air exchanges/hr.

A

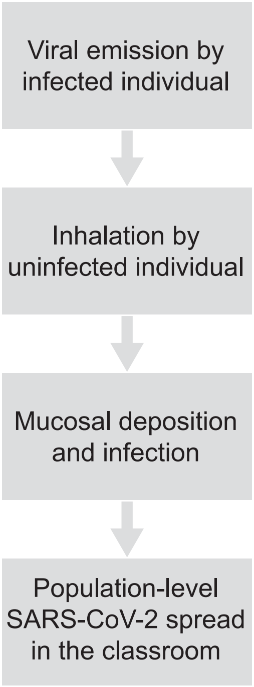


B

| Parameter name | Value |
| --- | --- |
| Classroom size | 4000 cubic feet |
| Low viral load emission rate | 5000 virions/hr |
| Intermediate viral load emission rate | 4,307,619 virions/hr |
| High viral load emission rate | 5,000,000 virions/hr |
| Viral particle lifetime | 1.6 hr |
| # of virions needed to initiate infection | 500 |
| Respiratory tidal volume | 0.5 L |
| Respiratory rate | 15 breaths/min |

**Figure S2:** Modeling approach to estimate transmission risk in the classroom. **A:** Model schematic. **B:** Table of parameter values used in differential equations model simulating SARS-CoV-2 emission and inhalation in classrooms.

### Supplementary Tables

| Droplet size (μm) | Percent of expelled droplets | Deposition probability |
|---|---|---|
| 0.10 | 0.16 | 1.34 |
| 0.20 | 0.16 | 1.60 |
| 0.30 | 0.16 | 1.53 |
| 0.40 | 0.16 | 1.51 |
| 0.50 | 0.16 | 1.47 |
| 0.60 | 0.16 | 1.47 |
| 0.70 | 0.16 | 1.52 |
| 0.80 | 0.16 | 1.52 |
| 0.90 | 0.16 | 1.48 |
| 1.00 | 0.16 | 1.43 |
| 1.50 | 0.16 | 1.74 |
| 2.00 | 0.16 | 1.98 |
| 2.50 | 0.16 | 2.49 |
| 3.00 | 0.16 | 2.82 |
| 3.50 | 0.16 | 3.70 |
| 4.00 | 0.16 | 4.77 |
| 4.50 | 0.16 | 5.61 |
| 5.00 | 2.44 | 5.97 |
| 6.00 | 2.44 | 6.71 |
| 7.00 | 2.44 | 6.09 |
| 8.00 | 2.44 | 5.36 |
| 9.00 | 2.44 | 4.69 |
| 10.00 | 1.70 | 4.07 |
| 11.00 | 1.70 | 3.54 |
| 12.00 | 1.70 | 3.09 |
| 13.00 | 1.70 | 2.94 |
| 14.00 | 1.70 | 2.73 |
| 15.00 | 0.90 | 2.18 |
| 16.00 | 0.90 | 1.55 |
| 17.00 | 0.90 | 1.23 |
| 18.00 | 0.90 | 0.78 |
| 19.00 | 0.90 | 0.73 |
| 20.00 | 0.78 | 0.52 |
| 21.00 | 0.78 | 0.41 |
| 22.00 | 0.78 | 0.29 |
| 23.00 | 0.78 | 0.17 |
| 24.00 | 0.78 | 0.07 |
| 25.00 | 0.84 | 0.01 |
| 26.00 | 0.84 | 0.00 |
| 27.00 | 0.84 | 0.01 |
| 28.00 | 0.84 | 0.00 |
| 29.00 | 0.84 | 0.00 |
| 30.00 | 0.88 | 0.00 |

**Table S1:** Estimating the fraction of inhaled viruses that are deposited in the nasopharynx based on CFD results.
